# Development and piloting of a primary school-based salt reduction programme: formative work and a process evaluation in rural and urban Malawi

**DOI:** 10.1101/2022.07.20.22277598

**Authors:** Nozgechi Phiri, Yvonne Cunningham, Stefan Witek-Mcmanus, McDonald Chabwera, Shekinah Munthali, Jones Masiye, Albert Saka, Miryam Katundulu, Caroline Chiphinga Mwale, Dalitso Dembo Kang’ombe, Joseph Kamangila, Amelia C Crampin, Frances S Mair

**Author notes:** Corresponding author (NP).

## Abstract

**Introduction:** Excess salt intake is a major modifiable risk factor for cardiovascular disease. Promoting salt reduction as part of routine school-health programming may be a pragmatic way to address this risk factor early in the life course but has not been tested in sub-Saharan Africa (SSA). Here we describe the formative work with stakeholders and process evaluation of pilot work to develop a school-based salt reduction programme for children aged 11-14 years, in preparation for a cluster-randomised trial in rural/urban Malawi.

**Materials and methods:** Collection of observational data and documentary evidence (meeting minutes/field notes) from the earliest key stakeholder engagement with Malawi Ministries of Health, Education, Local Government and Rural Development and Malawi Institute of Education, and non-governmental stakeholders; and a series of semi-structured interviews and focus groups (with head teachers (n=2); teachers (n=4); parents (n=30); and learners (n=40)). Data analysed thematically and conceptualised through a Normalization Process Theory lens.

**Results:** Formative work illustrated a range of administrative, technical, and practical issues faced during development of the programme; including allocation of stakeholder roles and responsibilities, harmonisation with pre-existing strategies and competing priorities, resources required for programme development, and design of effective teaching materials. While participants were positive about the programme, the process evaluation identified features to be refined including perceived challenges to participation, recommended adaptations to the content and delivery of lessons, and concerns related to quantity/quality of learning resources provided.

**Conclusion:** This study demonstrates the importance of comprehensive, sustained, and participatory stakeholder engagement in the development of a novel school health programme in SSA; and highlights the factors that were critical to successfully achieving this. We also demonstrate the value of detailed process evaluation in informing development of the programme to ensure that it was feasible and relevant to the context prior to evaluation through a cluster-randomised trial.

## Introduction

Non-communicable diseases (NCDs) are a growing public health concern in Low and Middle Income Countries (LMICs) especially in sub-Saharan Africa (SSA).[1, 2] Hypertension is the leading and most important risk factor for cardiovascular diseases (CVDs), which are a major cause of death.[3] There is therefore a growing need for population-based interventions.

An important modifiable risk factor for hypertension is sodium intake,[4] which is predominantly related to the level of salt in diets.[5] The World Health Organization (WHO) has set recommended maximum daily intakes; more than 2g per day of sodium intake (equivalent to 5g of salt) risks development of hypertension.[6-8] Globally, estimated mean sodium consumption is 3.95g, equivalent to 9.88g of salt per day.[9] Although previously the highest sodium intake was seen in high income countries,[9] similar trends are now evident in SSA. While specific statistics about salt intake in children in SSA are largely unavailable,[10] higher sodium intake in urban rather than rural areas of SSA has been reported [11] among both adults and children.[7, 12, 13] There is emerging evidence that dietary salt reduction can reduce blood pressure.[5, 7] A meta-analysis of 10 trials in participants aged 8-16 years showed that a 4-week modest salt intake reduced mean blood pressure by 1.2/1.3mmHg.[6, 14, 15]

The WHO’s global action plan for the prevention and control of NCDs 2013-2020 stipulates a relative 30% reduction in salt intake as one of the ‘best buys’ of the nine global NCD targets.[16] Malawi has a high discretionary salt intake and therefore individual or household salt reduction has the potential to be a highly cost-effective strategy for reducing hypertension and cardiovascular diseases.[2] This evidence provides the rationale for the proposed trial of a salt reduction intervention in schools in Malawi.

The ‘No to Sodium’ (No to Na) trial aims to evaluate a 12-week salt reduction programme embedded in the science and technology curriculum of primary school children (aged 11-14) through a cluster-randomized trial in rural (Karonga district) and urban (Lilongwe district) Malawi. This trial builds on a previous trial evaluating the effectiveness of schools in reducing the salt intake of children and their parents.[6, 15] In this article we describe formative work done through a process of stakeholder engagement, lessons learnt from a pilot of the programme and how these influenced the design of the final programme to be evaluated in the main trial. We used Normalization Process Theory (NPT) as our underpinning theoretical framework. NPT has been used extensively for designing and evaluating complex health interventions[17] and has been shown to help trialists ‘describe, assess and enhance implementation potential’.[17, 18] Our aims were:

1. To describe the formative work of stakeholder engagement undertaken to develop and refine a salt reduction programme in Malawian primary schools.
2. To describe qualitative findings relating to perceptions of the programme, including identified challenges and facilitators likely to influence success of implementation of the programme during the trial.

## Materials and methods

### Design

We describe the qualitative data collection methods employed to address our research aims which involved formative work (Phase 1); and a process evaluation of the programme pilot (Phase 2).

### Planned trial

This study was conducted in preparation for evaluation of the programme through a cluster randomized intervention trial in 11–14-year-old school attenders and their families in rural and urban Malawi. The trial involves 26 schools, with thirteen delivering the 12-week programme (consisting of lessons and competitions for children and other activities with parents) on salt reduction to children and their family members as part of the routine curriculum, and thirteen control schools following the routine curriculum only. Full details of the trial are described elsewhere. (https://doi.org/10.1186/ISRCTN13909759)

## Ethics statement

Ethical approval was received from the National Committee on Research in the Social Sciences and Humanities in Malawi (P.11/18/333) and the College of Medical, Veterinary and Life Sciences Ethics Committee at the University of Glasgow. Verbal and written information about the study were provided, in participants’ mother tongue prior to obtaining written informed consent. All Focus Group Discussions (FGDs) with adolescents only took place after obtaining written signed consent from parents and written signed assent from adolescents. This study is registered at ISRCTN (#13909759).

### Data collection

#### Phase 1 Formative work (preliminary engagement activities undertaken to develop and refine a school-based salt reduction programme)

The formative work included engaging with local stakeholders in key ministries: Ministries of Health (MoH), Education (MoE), Local Government and Rural Development (MLGRD) and the Malawi Institute of Education (MIE), and non-governmental stakeholders (from public and private sector including the business community) and adolescents and their guardians, to assess the feasibility and acceptability of the programme before implementation of the trial (Figure 1).

**Fig 1.**
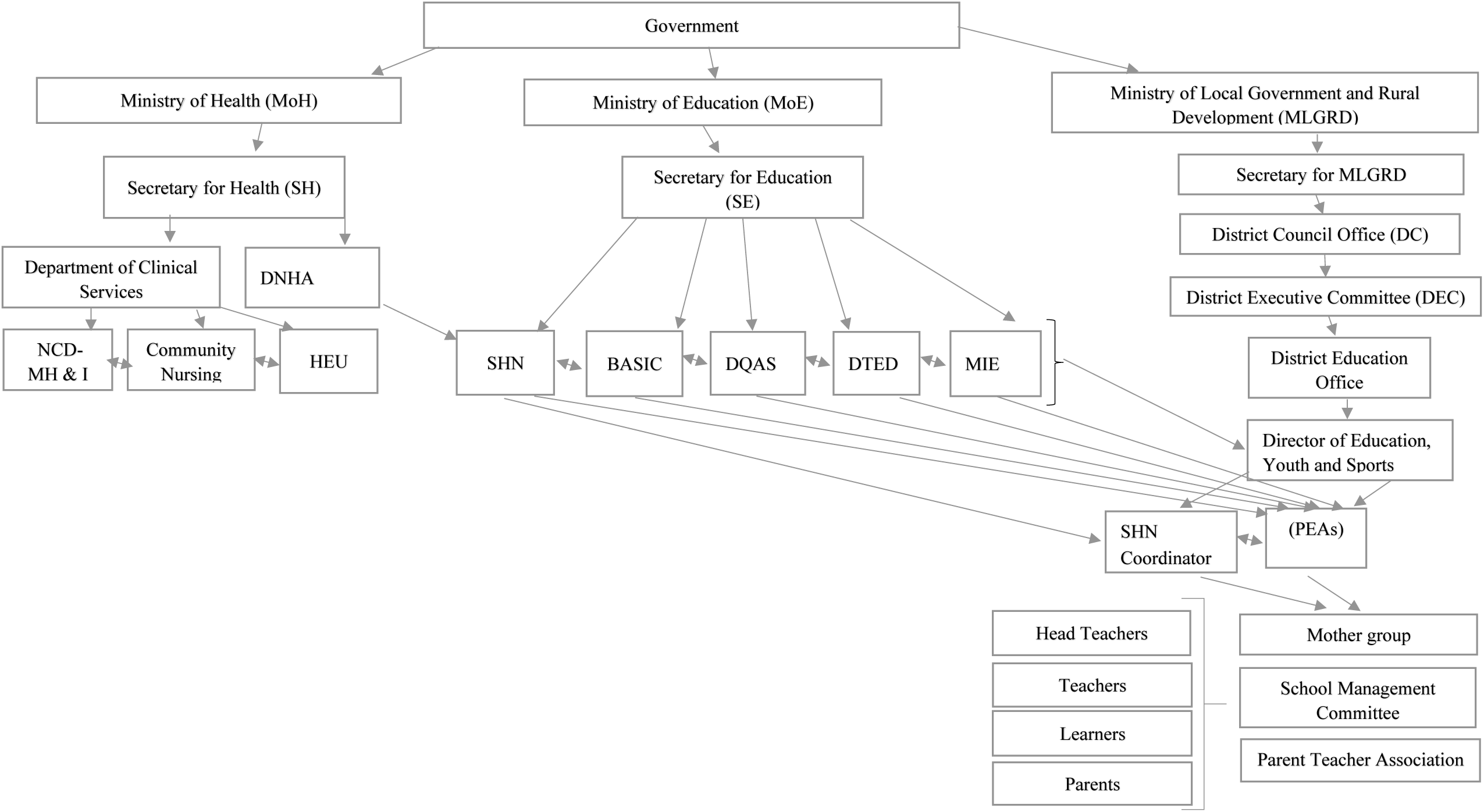
A pictorial chart of key stakeholders.

The School Health and Nutrition (SHN) Programme, a joint programme between MoH and MoE, had been engaged at grant proposal stage (through individual meetings and e-mails) and had negotiated support letters for the application from MoH and MoE. On award of the funding, further meetings took place with the ministries to generate an action plan for stakeholder engagement. The process of stakeholder engagement continued with a series of small meetings between the study leads and key people in the MoH and MoE (Fig 1). These meetings involved small groups (less than 5) and were held in their offices. The study team helped stakeholders understand the proposed trial by making comprehensive power point presentations at the beginning of each meeting. The meetings explored views on the planned trial and provided the opportunity to ask questions, enabling the stakeholders to get a clear understanding of the trial and its objectives.

Data was collected through minutes of meetings and field notes, from the very earliest stages of engagement which documented the extensive preliminary work undertaken (Table 1) from when the outline proposal was developed through to when funding was received, and materials were developed. This initial evaluation work explored how key stakeholders viewed the proposed programme, and factors influencing these perspectives and responses.

**Table 1.** Showing stakeholder engagement meetings.

| **TIMELINE** | **ACTIVITY** |
| --- | --- |
|  | **FIRST APPLICATION** |
| 8<sup>th</sup> December 2015 | PI meeting with MoH Non- Communicable Diseases, mental health, and Injury (NCD- MH & I) department lead regarding application |
| 12<sup>th</sup> December 2015 | PI introduced proposed study to SHN leads through NCD-MH & I department lead at MoH (co-applicant) |
| December 2015- January 2016 | Calls and informal meetings with MoH and SNH leads on study |
| 20<sup>th</sup> January 2016 | Letter of Support MoH, Letter of Support MoE through SHN |
| 21<sup>st</sup> January 2016 | Outline proposal submission |
|  | **Outline proposal rejected** |
|  | **SECOND APPLICATION** |
| May-June 2017 | Calls and informal meetings MoH and SHN leads |
| 15<sup>th</sup> June 2017 | New outline proposal submitted with original MOH- MOE letters of support and addition of new head of NCD-MH & I Department at MoH |
|  | **Outline proposal approved** |
| Aug-Sep 2017 | Calls and informal meetings MoH and SHN leads |
| 12<sup>th</sup> Sept 2017 | Revised Letter of Support from MoH and MoE through SHN |
| 20<sup>th</sup> Sept 2017 | Full Grant application Submission |
| 1<sup>st</sup> November 2017 | **Full proposal approved** |
|  | **ACTIVITY ON GRANT** |
| 11<sup>th</sup> June 2018 | Approval Letter |
| 8<sup>th</sup> November 2018 | MoH-MoE Stakeholder meeting- Discussed name of trial, need to decide focal ministry, iodine reduction, other stakeholders to include |
| 7<sup>th</sup> December 2018 | MoE Stakeholder meeting at MoE offices- to rope in relevant departments and affiliates- |
| 8<sup>th</sup> Feb 2019 | MoH, MoE Stakeholder meeting- included other stakeholders (university, NGOs and business entities) |
| April 2019 | 1) Department of Nutrition and HIV/AIDS (DNHA), NCD- MH & I and SHN (MoH)- discussion on lead department-<br>2) District Commission (Lilongwe and Karonga)- Gave approval to approach District Executive Committee (DEC), District Education Office and other relevant stakeholders (Primary Education Advisors-PEAs, SHN coordinator)<br>3) Directorate of Quality Assurance and Standards (DQAS)- Discussed the possibility of adding lessons as part of the curriculum. |
| April- May 2019 | Translation of curriculum and drawing of illustrations |
| 02<sup>nd</sup> May 2019 | Presentation to Karonga District Management Team and vote of approval |
| 3<sup>rd</sup> May 2019 | Presentation to Lilongwe City council and vote of approval |
| 06-08<sup>th</sup> May 2019 | Curriculum development workshop (DNHA, SHN, NCD-MH & I, DQAS, MIE, BASIC) |
| May 2019 | Editing and formatting of curriculum to match Malawi primary school curriculum by MIE and approved through DQAS. |
| May- June 2019 | Meetings with head teachers, Parent teacher associations (PTAs) and School Management committees (SMC) |
| June 2019 | Teacher's training |
| June-July 2019 | Pilot trial |
| 09-11<sup>th</sup> September 2019 | Curriculum refinement workshop (DNHA, SHN, NCD- MH &I, DQAS, MIE, BASIC) |

This preliminary work included developing and adapting the salt reduction programme using the content of the School-EduSalt Trial conducted in Changzhi, northern China[6] as a starting point, to suit the needs of Malawian adolescents and fit with the existing primary science and technology education curriculum in which health education is integrated. This work included adapting content, illustrations, and food examples to suit the Malawian context. Prior to adaptation the materials were translated to English, and the elements being shared with the learners families, were translated after adaptation into the local languages (Chichewa/ Chitumbuka). Learning materials were edited and formatted to be consistent with other science topics during a stakeholder workshop with two MoH departments (Department of Nutrition and HIV/AIDS-DNHA & Non-Communicable Diseases, mental health and Injury NCD-MH & I), four MoE departments (School Health Nutrition-SHN, Directorate of Quality Assurance and Standards-DQAS, Basic Education (BASIC) Department and Department of Teacher Education and Development-DTED) and one Ministry of Education affiliate organization (MIE), (Fig 1).

#### Phase 2: Process evaluation of a programme pilot, including identification of barriers and facilitators to successful implementation

The pilot programme took place in two primary schools in urban Lilongwe and rural Karonga. Standard 6 teachers received three days of training that introduced the programme, the intervention, and associated resources (e.g., parent letters, posters) and activities (e.g., parent forum, quiz and artistic performances). All learners in “Standard 6” (sixth year of universal free primary education in Malawi) received the programme for 12 weeks. The programme included weekly lessons that covered salt and health, harmful effects of salt and guidance on salt reduction strategies. Four lesson summary letters (in local language) were sent home with learners for parents to read. Three of the letters asked the parents to respond to several simple questions from the summary as an assessment of their understanding of salt intake and hypertension, further engaging parents and indicating the number of actively participating parents. Learners also participated in after-school activities either on their own in artistic competitions (poems, drama, song performances) or with a parent during family quizzes. Parents participated in a quiz and forum discussing their experience with the programme.

Teachers purposefully selected and invited learners to Focus Group Discussions (FGDs). Children took home letters with an information sheet detailing the trial information and consent forms (in the local language) for parents and guardians, which they returned after being signed. We conducted four FGDs comprising ten learners each who had participated in the pilot (Table 2).

**Table 2.** Summary of phase 2 data collection methods and participant characteristics.

| Participant type | Data collection method | Number of interviews/ FGDs | Number of participants |
| --- | --- | --- | --- |
| Head teachers | Semi-structured interviews | 2 | 2 |
| Teachers | Group interviews | 2 | 4 |
| Parents | FGD | 3 (one FGD males/one females/one mixed gender) | 30 |
| Learners | FGD | 4 (two with boys, two with girls) | 40 |

A convenience sample of parents and guardians were invited to participate in FGDs via a letter and information sheet taken home by learners. Teachers were also invited to participate in FGDs which lasted 40-60 minutes. Semi-structured interviews were held with head teachers from each school.

A semi-structured interview guide was used to gain perspectives on the learners’ views of the programme, including what they liked or disliked and their suggestions for improvement. For parents, we explored their experiences with the programme, the learning from their children and their interactions with teachers and the research team through parent meetings. We conducted seven non-participatory observations of three lessons delivered to learners. Interviews and FGDs were audio recorded and transcribed verbatim with the transcripts serving as data for analysis.

### Data analysis

Phase 1 and 2 data (minutes, field notes, transcripts, and observations) were coded in NVivo 12 using a thematic approach to analysis as outlined by Braun and Clarke.[19] Transcripts were initially coded by three separate researchers to develop a codebook. These researchers discussed any disagreements in the coding through virtual meetings fortnightly to ensure consistency of coding. Once the coding frame was established, all data was double coded. Field notes, minutes and transcripts were analysed using a thematic approach to identify participants’ main concerns on the salt reduction education and their perspectives on the curriculum, delivery mechanisms, family and community involvement as well as policy implications. The identified themes were then mapped onto the four main constructs of the NPT theoretical framework; coherence (sense-making), cognitive participation (engagement work), collective action (operationalization work), and reflexive monitoring (appraisal)[18] to help conceptualize the data. Themes that fell outside the framework were noted but not excluded from analysis. In this way, we avoid “shoe-horning” of the data and help identify issues that might still be important but not fit with our theoretical model. Finally, a descriptive summary was written to capture the information (with exemplar quotes) in each broad theme. The notes from the formative work were summarized to form a description of the processes to inform implementation of the programme during the trial.

## Results

### Phase 1: Formative work

Issues addressed and resolved during stakeholder meetings are as outlined below.

#### 1. Leadership and departmental involvement

A key question was which Ministry would be considered the focal or secretariat Ministry for the trial. As a health-related trial embedded in schools, clarification was required whether it should be under Ministry of Health or Ministry of Education (Fig 1) and positively, both Ministries were keen to assume leadership. Nutrition related initiatives in Malawi are often spearheaded by the Department of Nutrition and HIV/AIDS (under the Ministry of Health). However, this trial was deemed unique because it involved delivery of the intervention as part of the science curriculum, hence the Ministry of Education felt the trial should be overseen by their Ministry. Even within the Ministry of Health, there was uncertainty whether the trial should be overseen by the department of Non-Communicable Diseases and Mental Health & Injuries (NCD-MH & I) or the Department of Nutrition and HIV/AIDS (DNHA). After internal discussions the Ministry of Health assumed the lead through the DNHA. Department of School Health and Nutrition (SHN) was mandated by DNHA to oversee trial implementation because of its links to both the Ministry of Health and Ministry of Education (Table 1 and Fig 1). School Health and Nutrition with NCD-MH & I worked with the study team during implementation of the study while DNHA was to be engaged at a minimum capacity, with a later role of translating results into nutrition policy. Despite reaching agreement there were residual concerns from other relevant government departments and SHN was instrumental in achieving final consensus and encouraging full participation.

Further smaller meetings provided an opportunity for the School Health and Nutrition Department to identify other key departments within the Ministry of Health and Education to be included in the next phase of meetings e.g. Department of Quality Assurance and Standards (DQAS) responsible for supervision and inspection of teaching and learning resources in schools, Basic Education (BASIC) responsible for teacher’s welfare, Health Education Unit (HEU) responsible for promoting health education and Malawi Institute of Education (MIE) an affiliate of the Ministry of Education mandated to design, develop, monitor and evaluate primary school and teachers curriculum.

#### 2. Impact on iodine fortification programme

There was a concern from the DNHA representatives, that the intervention might affect iodine intake, as salt in Malawi is fortified with iodine as a public health measure to prevent goitre. Presentations by the study team were made using Malawi data from the Public Health Institute demonstrating that children and adults have a consistent excess iodine intake in all districts of Malawi and highlighting the tension between salt advertising capitalizing on the iodization programme (e.g., “Salt is good for your health”) and picturing high levels of salt application to food.

Another concern was the study title: ‘No to Na’ that might suggest total cessation of salt intake and lead to inadequate iodine intake undermining the extensive work that had previously been undertaken to improve iodine intake over the years. The NCD-MH & I department highlighted that health care workers are already advocating for reduction of salt as management or treatment strategy for hypertension and other NCDs. It was agreed to use an alternative title during the trial, and in interactions at district and community level. The study title was “Healthy Diets Study” to reduce risk of disclosure of the intention of the behavioural intervention in the control schools while maintaining “No to Na” as the protocol title as submitted to Ethics Committee.

#### 3. Integration into science curriculum

The Ministry of Education through the Department of Quality Assurance and Standards (DQAS) expressed concerns regarding the proposal to integrate the intervention into the science curriculum. The DQAS was concerned that with the school curriculum for the year already set, the only option was to offer it as standalone lessons. DQAS reviewed the proposed lessons with the support of the Malawi Institute of Education (MIE) and approved them to be piloted as standalone lessons, one lesson at the end of the day, during standard school hours, once every week.

After resolving these issues with key ministry stakeholders, a kick-off meeting was held at the beginning of 2019 with fifteen attendees (Table 1). The meeting was held to introduce and discuss the trial with a larger and diverse stakeholder group.

Subsequent meetings were held at the district level of government with the district executive committee (DEC) in Lilongwe (32 attendees) and Karonga (53 attendees), technical committees that advises and supports the District Councils (Table 1). As the engagement continued other smaller office meetings were held with people directly involved in supervising the schools e.g. the District Education Office, Primary Education Advisors (PEA) and School Health and Nutrition coordinators.

#### 4. Resources required

Although extensive stakeholder engagement was crucial to ensure successful implementation of the trial overall, the trial team had not anticipated the number and size of meetings that would ultimately be required. Pre-application discussions had not identified the number of different Ministry departments that would need to be involved, nor the complexity of processes required to generate and authorise the materials to be used. This important formative work resulted in increased study costs to cover conference, subsistence, travel and accommodation costs and delayed the start of piloting. Stakeholder engagement began in September 2018 and the pilot took place in June-July 2019.

#### 5. Development of teaching materials

Before the pilot began, materials from the Edu-Salt programme[6, 15] were translated from Mandarin to English to generate a template. The materials were then significantly adapted to reflect the Malawian context, taking due note of the major difference between urban and rural diets and lifestyles. The study team originally proposed separate materials for urban and rural communities, but the Ministry of Education (MoE) was clear that if successful, in order to make the programme scalable only one set of materials should be used. The study team also proposed that all materials be in the local languages (Chichewa for urban Lilongwe, and Chitumbuka for rural Karonga). The MoE team were again clear that English was the official language of instruction for Science in Standard 6 and that lessons must be conducted in English with English materials. Concerns about this were addressed by the Education team who were clear that that the teachers may and do explain difficult words or extracts to learners in their local language to help them grasp the topic, then switch back to English. The exception that was negotiated was the materials to be shared with the parents (letters and flyer) which were to be prepared in the two relevant languages. A local illustrator was engaged to add illustrations to match the content in the curriculum (Fig 2).

**Fig 2.**
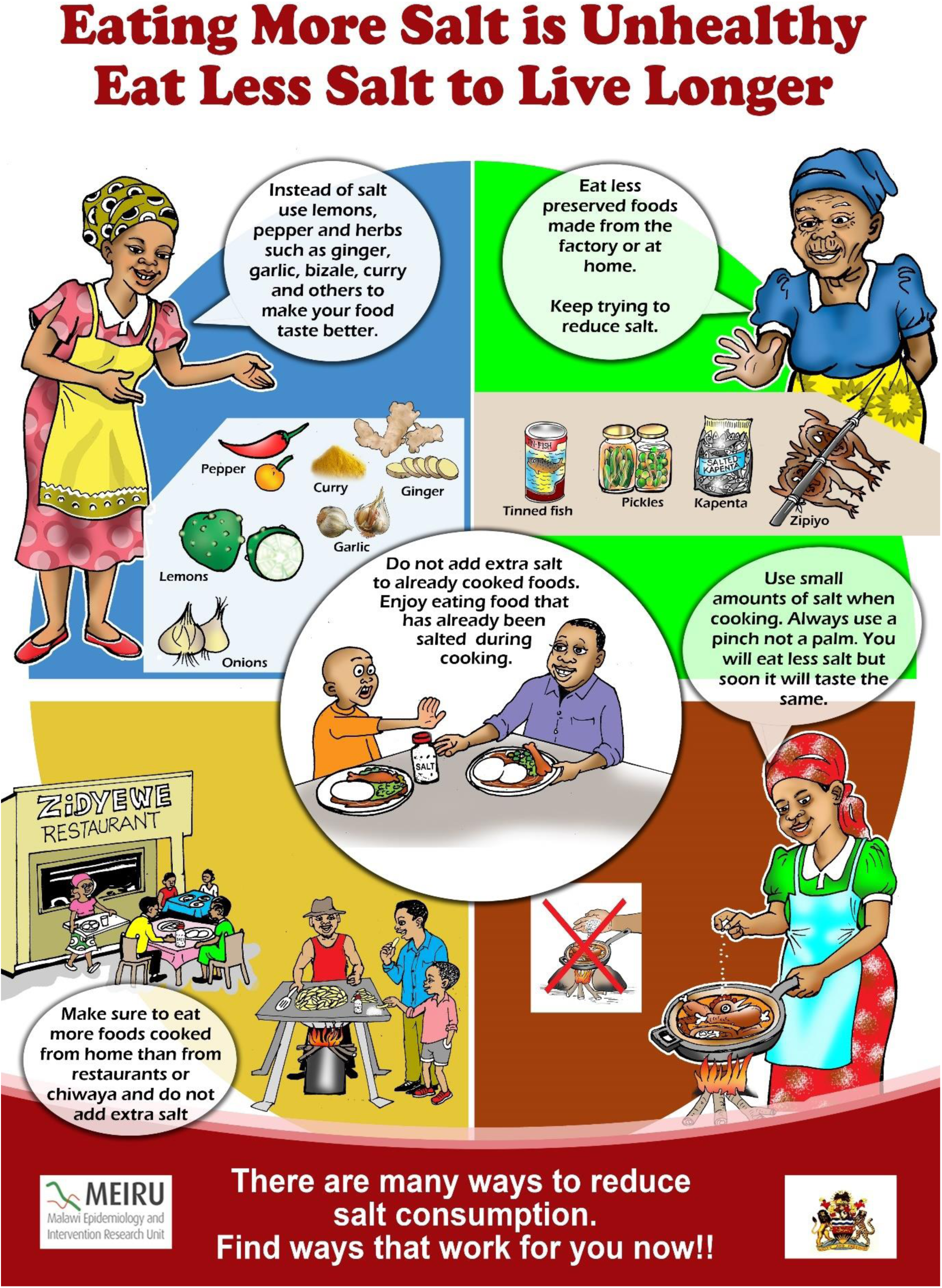
Flyer for parents showing strategies for reducing salt.

The materials and programme were discussed in detail at a programme development workshop that included the key Ministry departments and affiliates as well as teachers and head teachers (21 attendees). Following this workshop, the programme was further modified and tailored to fit the Malawian curriculum format.

### Phase 2: process evaluation of the programme pilot

The findings from the pilot of the programme are presented under five key themes: 1) Sense-making: teacher, parent and learner views of a school-based intervention for reducing salt (coherence); 2) Relationship work: engaging learners and through them parents in the salt reduction programme (cognitive participation); 3) Enacting work: teachers, parents and learners doing the work of participating in the programme (collective action); 4) Perceived facilitators to the programme (reflexive monitoring); 5) Perceived challenges to the programme (reflexive monitoring). These themes are described, and illustrative quotations provided.

#### 1. Sense-making: teacher, parent, and learner views of a school-based programme for reducing salt intake (coherence)

Learners and parents were positive about the programme and were able to identify possible adverse effects of a high-salt diet. Participants also recognised their households’ high salt intakes and spoke about how they were able to reduce this and become accustomed to the taste of less salty food. The learners’ FGDs demonstrated learning relating to each of the programme units including the harmful effects of excessive salt, though some had difficulties understanding the recommended daily salt intake and how best to measure and reduce their salt intake. Some parents also spoke of similar challenges as highlighted below:

> *The way I understood from the several meetings I have been. This is the second one. I heard that the whole household is supposed to eat less than 5 grams per day. So, you will correct me if am wrong right here [Interviewer 2: we need to correct each other] we should eat less than 5 grams of salt [Interviewer 1: Per person?] no per family. [Parent]*

Others spoke of putting their learning into practice; and some interviewees linked this new learning to their existing knowledge, for example about people in their family affected by relevant conditions such as high blood pressure as shown below:

> *“I: So why did you decide to tell your parents?*
>
> *P: To protect their life as my mother already has ulcers and my father has Blood Pressure, so we were taught that even our parents should not eat too much salt.” [Learners]*

#### 2. Relationship work: engaging learners and through them parents in the salt reduction programme (cognitive participation)

Learners told their extended families and neighbours about what they had learnt about salt reduction. This communication was supported by the letters from schools to parents.

> *When the child got home, he said “I have come with this letter from KPS [Karonga Prevention study now known as Malawi Epidemiology and Intervention Research Unit-MEIRU] who were teaching us at school” then I asked him, what did they teach you? He said, they gave us a letter and told us to give to our father, so here is the letter. I got the letter as an individual and started reading. When I finished the letter, I invited my family and at home I have a mother, my sister, two wives and children, then I started explaining. [Parents]*

Parents were invited to a parent forum and then a quiz. At the parent forum, parents discussed the work their children did in class and at the quiz, two groups of parents competed with one another, answering questions about salt reduction. Teachers thought engaging parents consolidated the information that learners delivered at home and meant parents could work through challenges to reducing salt intake. Parents valued these meetings as it allowed them to learn from other parents and these meetings motivated parents and learners to participate in the intervention as illustrated below.

> *“When we took part in the quiz, we were encouraged that we are not alone in this study seeing our fellow parents participating and even the children were encouraged seeing parents there, thank you very much.” [Parent]*

Some parents said they agreed to participate because of previous experiences with the research group, MEIRU which has shown them that their work is honest and aims to help people.

The parents who attended FGDs were generally very positive about the parent meetings, however only half the parents who were invited attended. Some parents explained that the non-attenders could not come as they worked or went to church on Sundays, but others maintained that they were negligent. Most parents said that providing incentives, such as drinks or snacks, would encourage more parents to attend meetings. Teachers also valued incentives as this quote shows.

> *And we felt when you were coming to our schools to do that exercise with learners, after classes we thought the learners will be given a little something whether Mahewu [maize drink] just to keep them up to the knocking time. [Head teacher]*

Parents’ suggestions for improving participation included: a) providing incentives/gifts to encourage parent attendance; b) sending meeting invitations well in advance to give parents time to prepare; c) sharing meeting minutes with absentee parents through their children so that they could see what they missed; d) engaging with the community through the village chiefs, where the intervention could be explained to everyone.

#### 3. Enacting work: teachers, parents and learners doing the work of participating in the programme (collective action)

Learners, parents, and teachers described the practical steps they took to reduce salt. Participants explained how they reduced their salt intake, some telling stories of, for example, planting garlic to provide an alternative flavouring or buying a spoon for measuring the salt. Others said they had stopped putting salt on the table or hid it from others in their house to ensure that they did not add extra salt to food.

> *“I have already explained about challenges which I face. My other family members complain whenever I add little salt and they add extra salt. So, I take that salt and hide it somewhere so that they should think that we have run out of salt.” [Parent]*

This behaviour change met challenges. Parents had difficulties in reducing salt intake as some family members (or visitors) resisted, as they found it difficult to become accustomed to the new taste as this parent explains.

> *“…the challenge which I faced was we were separating relish because my husband was refusing to take low salt. He was saying he can’t manage to follow this advice because he is accustomed to taking more salt and he can’t start taking little salt. He also said, this is unhealthy because a person can’t eat relish which he/she feels does not taste good.” [Parent]*

Learners also reported some resistance to reducing salt intake from relatives. Some learners talked about resistance from their parents who did not want to listen to their children and who did not want to reduce their salt intake. This learner explains:

> *“…when I am telling them they say, “it’s just a topic which you are just learning at school so that your future should be bright” so it’s what they say.” [Learner]*

Some parents agreed that in the beginning it was difficult for them to listen to what their children were saying (though they did eventually take up the intervention).

#### 4. Perceived facilitators to the programme (reflexive monitoring)

There was positive feedback from learners, parents, and teachers about the programme and they reported the positive impact that the programme had in their lives. Some felt that their reduced salt intake had significantly improved their health, as this learner explains:

> *“I was one of the people who liked to add salt to foods that was already salted whether enough or a lot, I just had to add so I can taste saltiness. But after I have learnt I know that it’s good not to add salt to food that is already salted. It has helped my life.” [Learner]*

Teachers felt the programme was beneficial to the nation as the information gained by learners would spread to parents, neighbours, and the community. A teacher explains:

> *“…the project was a good one in a sense that we have learnt a lot from it. One, how to control our diet, how to control our sugar levels, how to control our salt levels and how we can manage ourselves. And most of all, learners have learnt a lot from it, in a sense that they have benefited something from it… So, we feel the project has a great impact to the nation because once a learner is taught, he will still be imparting that knowledge to the community, to the family members and to their friends as well. [Teacher]*

According to the parents’ FGDs, potential improvements to their lives, and crucially to their children’s lives, was the greatest motivation for engaging with the programme. Parents spoke of how their children were the future, and that the children liked to see their parents participating in meetings at the school.

Parents and teachers had suggestions about future expansion of the programme. They wanted the programme to be incorporated into the syllabus and expanded to the whole of Malawi but also to include other classes, as it was a programme which could benefit many learners and families.

> *“I just want to say you should continue with the program because a lot of people don’t know about salt, so reach out even in villages reach out to them, some don’t know that too much salt intake is harmful, even here in town, some people don’t know about salt.” [Parent]*

#### 5. Appraisal work: perceived challenges to the intervention (reflexive monitoring)

While teachers felt the programme was valuable, they noted some challenges, for example, the terminology used in the learning materials was unfamiliar to learners and made lessons difficult. Teachers and learners also mentioned language as a challenge as the learners had just transitioned from using Chichewa as a language of instruction to English in Standard 5.

> *“…of course, in class we have active learners, and we also have learners that learn at a slower pace, so we tried so that these slower pace learners should understand because our aim was the learners must teach the parents in the community and we could even try to translate the content from English to their local language for them to understand and take the message to their parent. [Teacher]*

Some of the challenges that teachers faced were a lack of time to complete the programme (some lessons had to be held outside normal school hours) and a lack of teaching materials (or unsuitable materials).

> *“…the only problem we had was on time; the time was really short, and I can say the content was over planned, but we tried our best to take time to teach and mainly the last period we used it to extend the period with 30 minutes for us to finish the lesson.” [Teacher]*

Individual schools and teachers also needed to supply some materials, such as ingredients to cook porridge (for a class experiment), this was difficult as the programme started midway through the school year when budgets had already been allocated but the ministry of Education had made it clear that teaching aids are prepared by either the school or the teacher.

> *“…the challenges that we met, mainly we had inadequate teaching and learning resources. Like we had to have packets of different types of salt. We are talking of kitchen salt, table salt which we can distribute in groups so that every group should have a look at the packet of salt which is not open. So, to get those, it was really a problem. [Teacher]*

## Discussion

This paper presents: 1) a description of extensive formative work undertaken with stakeholders to develop and refine a school-based salt reduction programme in preparation for a cluster randomised trial; and 2) the findings of the process evaluation undertaken during a pilot of the programme.

Stakeholder engagement preceding trial implementation in a LMIC is rarely reported and based on the findings presented here it is clear that it is time consuming and requires substantial investment. However, this effort was essential to obtain the support of key stakeholders, avoid misconceptions and allay any concerns, as well as to clarify which stakeholders would be responsible for implementation of the programme. This work reaped benefits as outlined in Table 3 and facilitated approvals for trial activities and minimised potential delays.

**Table 3.** Summary of key achievements and implications of the formative work.

| Key achievements for formative work | Impact |
| --- | --- |
| Agreed on leadership and key contacts for government departments and their roles in the deployment of the salt reduction programme in schools. | 1) Provided a sense of ownership and helped achieve “buy in” of key stakeholders.<br>2) Improved flow of information, minimised approval delays and resolved conflicts. |
| Addressed concerns about the trial with key government ministries and departments (Fig 1) and reached a shared understanding of the aims and methods of the trial. | Removed barriers to deployment of the intervention (for example, by alleviating concerns that the intervention might reduce iodine consumption or stop salt consumption completely and undermine previous public health messaging.) |
| Enabled identification and inclusion of relevant and diverse group of stakeholders. | The diversity of input optimised development and refinement of the intervention materials. |

The process evaluation of the programme pilot then identified aspects of the programme that needed to be refined or altered, for example, in relation to timing/duration of the lessons, content of the lessons and teaching resources, to improve the likelihood of successful implementation of the programme (see Table 4). Delivery of the programme was facilitated by the ease with which it could be implemented with minimal disruption to an already packed curriculum.

**Table 4.**
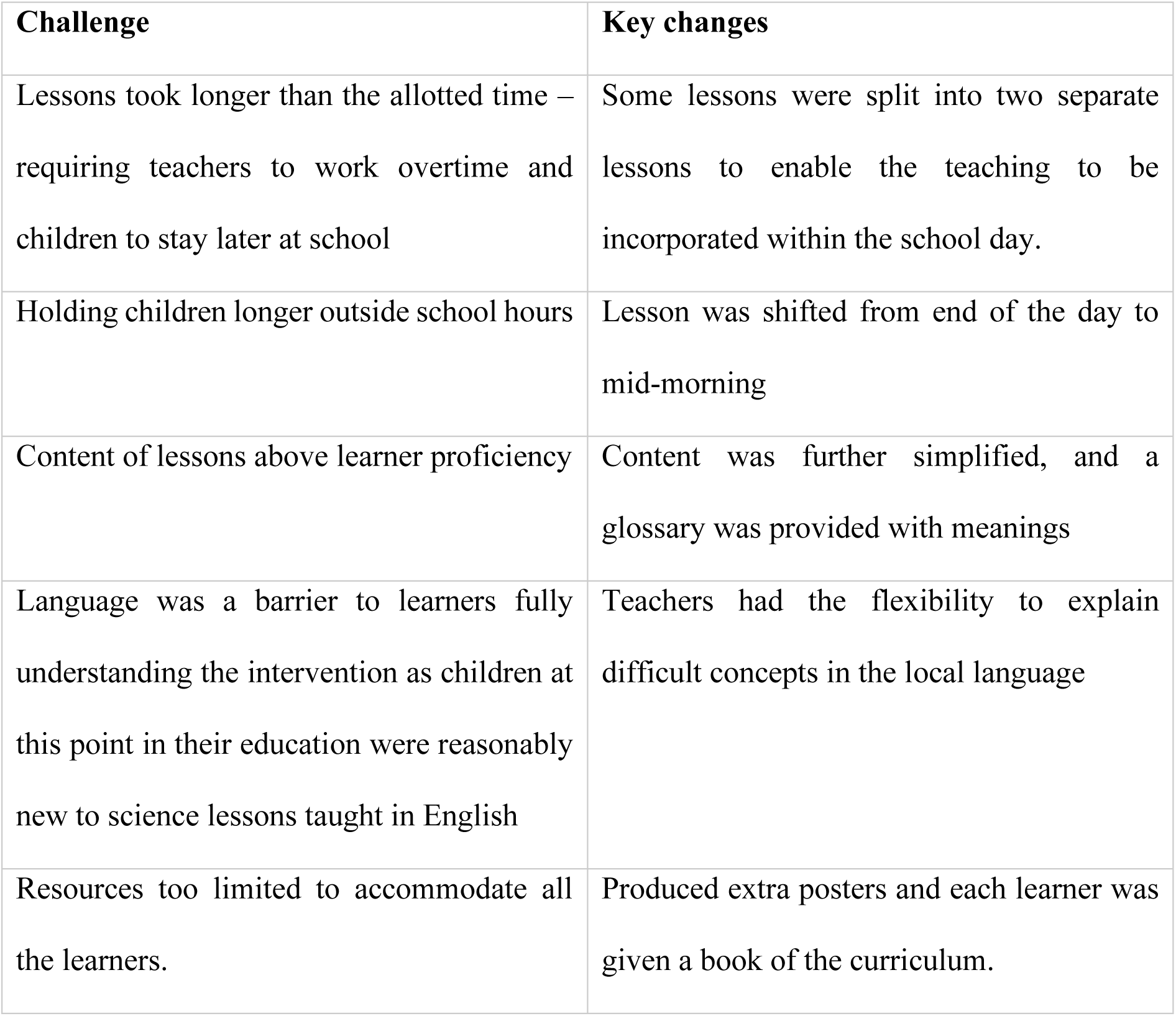
Summary of challenges identified in the programme pilot and implications for the trial.

The process evaluation of the programme pilot showed that parents, children, and teachers were generally positive about the programme. Students had the opportunity to contribute to the learning process by sharing their views on the topics.

Previous research has suggested that prevention programmes are most effective if the approach is multilevel, involving the learner, school and family.[20] Similarly, a report published by WHO,[21] concluded that the most effective school interventions are multicomponent and should include a curriculum taught by trained teachers, supportive school policies, a physical activity programme and healthy food served by the school canteen services (which rarely exist in the local setting). In this salt reduction programme participation in parent forums or quizzes was suboptimal and participants believed this was due to lack of incentives (e.g. snacks, money), which although they could have been provided in a trial setting would have made the intervention unsustainable. However, parents enjoyed parent forums as it showed them that they were not alone in their experiences with the programme and they believed their participation encouraged their children to comply with the programme.

Over the past two decades, school-based health programmes have become a key component of public health strategies in LMICs. For example, they have been used in the Middle East, Europe, Asia, Africa, and South America to promote adolescent mental health,[22] and in Zambia, South Africa, Tanzania and Belize to promote AIDS awareness and sexual and reproductive health.[23]

School-based health programmes focusing on diet and nutrition have included standards placed on the composition of school meals and limits to food choices available in school canteens as well as changes to school curricula to improve nutrition education.[21] School-based nutrition and diet interventions have been widely used with a recent umbrella review identifying thirteen separate systematic reviews describing eighty-two studies undertaken in the United States, Mexico, Canada, Europe, Asia, South America, the Middle East and Australia.[24]

A review of childhood obesity prevention interventions in Africa,[25] included 17 articles describing 14 interventions in three countries (South Africa, Tunisia and Uganda). This review identified barriers to implementing school-based interventions as largely resource-related, including lack of time from teachers and stakeholders, buy-in, training or motivation. Some interventions in China[26-28] (which were not focused on salt) involved parents and Wang et al.’s[29] study of a school-based initiative to reduce obesity noted that interactive and innovative intervention components facilitated the intervention.

Apart from No to Na, the only other interventions focused solely on salt reduction were the Edu-salt programme in China,[6] on which the No to Na intervention is based and the COMBI-ELS intervention in Vietman.[30] The COMBI-ELS intervention involved several components including targeted primary school interventions, mass media communications, training for cooks, and community programmes. It was not possible to ascertain which elements of the programme were most effective in changing behaviour nor was it possible to assess the contribution of the work in schools to the programme’s success.

A recent systematic review[31] examining the effectiveness of school-based nutrition interventions in sub-Saharan Africa found 14 studies based in South Africa, Botswana, Burkina Faso, Kenya, Nigeria, and Tanzania, but none focused on salt reduction, instead they focused on improving diets to reduce obesity and increasing physical activity. These studies showed that there was an improvement in nutrition knowledge following the interventions, but this did not necessarily translate into healthy nutrition behaviour. Just one school based intervention in South America (Trinidad and Tobago) included salt-reduction in its focus (in this case reducing the consumption of snack foods high in fat, sugar and salt) as part of a wider programme.[32]

### Strengths and limitations

A key strength of this study is that it describes the formative work undertaken to develop the programme and gives an account of the extensive engagement work required. The authors have not found any other reports of such work, though it is essential to inform future research and policy. Rigorous thematic analysis was conducted, drawing on both inductive (i.e., data-driven) and deductive (i.e., based on pre-conceived ideas) approaches, until data saturation was reached, and our analysis was underpinned by a robust theoretical framework, Normalization Process Theory. Double coding of all pilot process evaluation interviews and coding clinics enhanced reliability. However, the work was undertaken in Malawi and the findings cannot necessarily be generalised to other LMICs. We did not interview those who did not engage with the programme and so respondents may provide a more positive view of the intervention than their peers who did not engage.

## Conclusions

This study enhances our understanding of how a range of health, nutrition and education stakeholders view, experience, and value a school-based programme for salt reduction, and of the factors that may influence implementation and impact of the programme in a LMIC. Our work has highlighted the importance of extensive, preliminary engagement with a wide range of stakeholders to achieve “buy in” and active cooperation but has also demonstrated that future studies of this kind need to allocate substantial time and funds to this task. Our process evaluation identified five key challenges that led to refinements to how the programme was delivered. Our parallel process evaluation of the programme as planned to be implemented during the trial will help us understand whether these refinements helped and what further changes, if any, would be required to facilitate scale up beyond the trial or implementation in similar settings. Work such as this, which generates insights about the feasibility, acceptability, usability, and up-scalability of school-based programmes, is crucial for practice and policy decision-making on the future provision of novel health programmes in school settings.

## Data Availability

All data used in this study is available on reasonable request from Malawi Epidemiology and Intervention Research Unit. Please contact the Unit's Data Documentarist at

## Acknowledgements

We would like to thank all the participants who participated in the intervention and in the focus group discussions or interviews. We owe the head teachers, teachers, learners, and their guardians from the two primary schools our gratitude for their time, patience, and contributions. We also acknowledge and thank all key stakeholders who participated and contributed to the formative work. We would like to thank Hazel Namadingo and Cecilia Nyirenda for all their work and effort in this study. Gratitude should also go to James Kazembe for the tremendous work in the illustrations used in the intervention materials.

